# Radiation sensitivity and efficacy in aggressive and non-aggressive basal cell carcinoma (BCC) of the skin: Image Guided Superficial Radiation Therapy achieves high rate of local control in sclerosing, infiltrative, morpheaform and micronodular BCC subtypes as well as in non high risk BCCs, an analysis of 7994 BCC lesions

**DOI:** 10.1101/2024.07.17.24310584

**Authors:** Lio Yu, Michael Kaczmarski, Clay Cockerell

## Abstract

**Background:** High risk (HR) basal cell carcinoma (BCC) subtypes have been associated with high recurrence rates that is felt to be better managed surgically. Specifically, Mohs Micrographic Surgery (MMS) is considered most effective for aggressive HR BCCs and superior to traditional nonsurgical techniques, including radiation. Recently, superficial radiation therapy with high resolution ultrasound image guidance called Image Guided Superficial Radiation Therapy (IGSRT) displayed high local control (LC) rates and is an emerging non-surgical alternative to MMS for non-melanoma skin cancer (NMSC).

**Objectives:** We present the largest experience in the USA on treatment of BCCs using IGSRT and specifically evaluate if there are differences in LC between HR BCC versus non-HR subtypes using this technology.

**Methods:** A retrospective analysis was conducted on 7,994 BCC lesions treated with IGSRT in the continental United States. We compared the results of BCCs treated with IGSRT separated by HR vs non HR groups including 339 HR BCC lesions and 7655 non HR BCC lesions. High risk was defined as infiltrative, micronodular, morpheaform, and sclerosing subtypes. Non-HR BCC included superficial, nodular, and not otherwise specified (NOS) subtypes. Local control (LC) rates at two and five years were calculated with actuarial life-table and Kaplan-Meier methods and statistically compared using log rank tests.

**Results:** IGSRT treatment of the HR BCC group showed no recurrences with two and five-year actuarial and KM LC rates all at 100%. In comparison, the non-HR BCC cohort achieved similar two and five-year actuarial LC rates of 99.71% and 99.24% (KM LC at 99.5% and 99.23%), respectively. No statistical differences in LC rates between the two cohorts (p=0.278 each) resulted. Patients tolerated treatment well with little or rare high grade RTOG toxicity reported in both cohorts.

**Conclusion:** HR BCC may be treated just as effectively as low risk BCC using IGSRT and presents a viable alternative to MMS. The targeted approach using IGSRT, incorporating high resolution dermal ultrasound (HRDUS), appear to enhance treatment accuracy and effectiveness demonstrating high LC rates in all subtypes of BCC comparable to MMS and is a viable non-surgical option.

**Plain language summary:** *Effectiveness of a non-surgical skin cancer treatment using an image guided form of radiation modality on all subtypes of basal cell skin cancer:* Recent studies using a non-surgical treatment combining low penetrance radiation with ultrasound called Image Guided Superficial Radiation Therapy (IGSRT) showed promise in curing Basal Cell Cancer (BCC) of the skin, which is the most common skin cancer worldwide afflicting millions annually. Recent studies on early stage (I, II) BCCs treated with IGSRT (estimated combined total of ∼1900 BCC cases) appear to rival the best surgical treatment available called Mohs Micrographic Surgery (“Mohs” or MMS). Furthermore, certain subtypes of BCC appear to behave more aggressively with worse outcomes even with surgery and is generally felt inappropriate for radiation treatment. However, BCC subtypes were not specified in previous IGSRT studies. This study presents the largest experience (using medical chart review) in approximately 8000 BCC cases treated by IGSRT across the continental United States separated by aggressive vs non-aggressive subtypes for early stages (I, II) as well as more advanced (stage III) BCC cases to evaluate the efficacy and safety. This study confirms the high cure/control rate and safety of IGSRT for all subtypes of BCC which appear equivalent with Mohs (although the study was not meant to be a head to head comparison of the 2 different modalities). Moreover, the aggressive types of BCC showed similar (if not marginally better) cure rates than the more common non-aggressive BCC subtypes. The potential benefits to patients from this study show there is now a clinically proven non-surgical treatment with the same effectiveness as surgery for the most common cancer on the planet.

**Key Points:**

- This study provides evidence that backs up using IGSRT as a viable treatment option to MMS for both high risk and non-high risk BCC cases, achieving similar local control rates for both groups.
- It highlights that high risk BCC is more sensitive to radiation therapies such as IGSRT than previously believed, challenging the conventional practice of surgical treatment.

## 1 Introduction

Skin cancer is the most common form of cancer globally and presents a significant health challenge with various subtypes and varying prognoses. Among the many kinds of cancers, basal cell carcinoma (BCC) emerges as e most common type, accounting for nearly 80% of non-melanoma skin cancers [1]. As the incidence of non-melanoma skin cancers (NMSC) continues to rise with an estimation of 3.5 million patients with 5.4 million lesions each year diagnosed in the United States as of 2012 and continues to rise, the need for developing effective strategies to manage the disease becomes more imperative, particularly in the context of BCC [2].

For early stage BCC, which is the most common skin cancer amongst patients, several approaches are available for these cases including surgical excision, radiotherapy, topical imiquimod, photodynamic therapy [3,4] and Mohs Micrographic Surgery (MMS). Nevertheless, after BCC treatments (surgical or non-surgical) there is an increased risk of future BCC developing with up to a 16% chance of the development of new BCCs within the first year after initial treatment and an expected 5 year incidence for BCC developing of 45% as compared to the general US white population [5] which demonstrates the scope of the problem.

Although basal cell carcinoma has a characteristic of slow progression, local invasiveness, and low metastatic potential, there exist histological subtypes that are considered “high risk” that have been observed to have a more aggressive nature and ability to recur [6] Certain histologies such as morphemic, sclerosing, infiltrative, and micro nodular subtype present a challenge to typical treatment practices that have led to favoring surgical interventions over alternative modalities [7].

According to the American Academy of Dermatology (AAD), surgical interventions with Mohs micrographic surgery are widely recognized as the gold standard of care for patients with morpheaform and sclerosing BCCs due to their ability to conserve healthy tissue and retain high cure rates [8]. Conventional wisdom in dermatological oncology has a long held the belief that high risk BCCs are less responsive to radiation therapy and less prone to recur with surgery. This belief has been rooted by the inherent biological aggressiveness of these histologies backed by prior observational data [10] Surgical management of high risk cases such as morpheaform BCC on the nose was observed to have an incomplete excision rate of 61.5% and at the ear of 50% [11]. Coupled with various concerns about the efficacy and safety aspects of radiation therapies, this has led to vigilant or cautionary approaches in treating these high risk BCC histologies. However, this established approach may not be suitable for all patients. Several factors such as disfigurement, the overall health of the patient, or the ability to tolerate surgery treatments may have an influence on treatment decisions. Thus, it is crucial to consider alternative and less invasive treatments in selected cases [9].

As radiation therapy techniques have advanced recently, the introduction of image-guided superficial radiation therapy (IGSRT) which is a combination of HRDUS with SRT has allowed for the reconsideration of methods employed on treating skin cancer. Specifically, with regard to early stage NMSC (including high risk BCCs), IGSRT, which is regarded as precision and targeted medicine, potentially mitigates limitations associated with conventional non image-guided traditional radiation therapy (such a external/electron beam radiotherapy [XRT] and superficial radiation therapy [SRT]) and has been shown to statistically improve outcomes on treatment of all major histologies of NMSC, such as BCC, SCC and SCCIS [12–15]. On this basis, it stands to reason that re-assessment of the previous notion of bypassing the option of radiotherapy in high risk subtypes of BCCs should be evaluated using this more modern method of radiotherapy incorporating image guidance intrinsically to assess to see if the prevailing sentiment of non-radiation approach still holds true.

We set out to evaluate the differences, if any, in the local control rate of aggressive high risk BCC subtypes in comparison to the non-aggressive BCC subtypes treated with the use of this newer technology of IGSRT.

## 2 Methods and methods

From 2016-2023, pathologic proven BCCs of stage I, II,III (AJCC 8^TH^ edition cancer staging with tumor stage T1, T2, and T3) treated w IGSRT at multiple institutions throughout the continental United States were retrospectively reviewed and analyzed. Only pure BCCs without combined or collision histologic components (ie, without any squamous cell carcinoma (SCC), squamous cell carcinoma in situ (SCCIS), keratoacanthoma (KA), other non BCC histologies) were selected. We separated the high risk (HR) BCC histology cohort from the non-HR BCCs, gathered the lesion characteristics, treatment parameters and analyzed the rate of recurrence deriving the lesion local control post IGSRT treatment. Lesion and patient characteristics were summarized descriptively. Actuarial (life-table) local control (LC) rate and Kaplan-Meier (KM) LC were calculated and plotted graphically. Comparison of the actuarial LC and KMLC between the HR BCC vs non-HR BCC cohorts were performed using log-rank statistical methods.

High risk histology are defined as Infiltrative, Micronodular, Morpheaform, and Sclerosing subtype of BCCs. Non-HR BCC histology included superficial, nodular, or not otherwise specified (NOS) subtypes.

This study has been determined to be exempt by□WCG-IRB□under 45 CFR § 46.104(d)(4), because the research involves the use of identifiable□information which is recorded by the investigator in such a manner that the identity of the human subjects cannot readily be ascertained directly or through identifiers linked to the subjects, the investigator does not contact the subjects, and the investigator will not re-identify subjects.

## 3 Results

Patient/Lesion characteristics:

Between 2016-2023, 339 high risk (HR) BCC and 7655 non-HR BCC lesions were treated with IGSRT.

Table 1 summarizes the patient age and gender revealing a slight predominance of males (55.8%) overall however, there were slightly more females proportionally in the HR group. Mean age of patients was 72.5 ranging from 19.7 to 104.5.

**Table 1.** Patient Characteristics Tables *.

\***AGE AT FIRST TREATMENT**
|  | **ALL** | **(min-max)** | **HR** | **(min-max)** | **non HR** | **(min-max)** |
| --- | --- | --- | --- | --- | --- | --- |
|  | n =5758 |  | n = 339 |  | n =5419 |  |
| **Mean Age** | 72.5 | (19.7-104.5) | 72.8 | (40.0-97.5) | 72.1 | (19.7-104.5) |

**GENDER**
|  | **ALL** | **HR** | **non HR** |
| --- | --- | --- | --- |
| **Male** | 3208 | 166 | 3042 |
| **Female** | 2545 | 173 | 2372 |
| **Total** | 5753 | 339 | 5414 |

\*Not all patient genders and ages were available

Table 2 demonstrates the anatomic distributions of NMSC lesions separated by risk groups. As expected, the most common sites of BCC lesions were on the head and neck(H&N). There were more HR BCCs occurring in the H&N region in comparison to the non-HR BCC cohort (90% vs 80%).

**Table 2.** Anatomic distribution of NMSC lesions.

|  | HR BCC | NON-HR BCC |
| --- | --- | --- |
| **Head and neck (H&N)** | **289/321 (90.03%)** | **5662/7106 (79.68%)** |
| *H&N sublocation* |  |  |
| Ear | 55/321 (17.13%) | 627/7106 (8.82%) |
| Cheek | 38/321 (11.84%) | 1143/7106 (16.08%) |
| Nose | 135/321 (42.06%) | 2145/7106 (30.19%) |
| Cutaneous lip | 9/321 (2.80%) | 249/7106 (3.50%) |
| Mucosal lip | 1/321 (0.31%) | 12/7106 (0.17%) |
| Forehead | 26/321 (8.09%) | 702/7106 (9.88%) |
| Temple | 8/321 (2.49%) | 233/7106 (3.28%) |
| Scalp | 11/321 (3.43%) | 209/7106 (2.94%) |
| Neck | 6/321 (1.87%) | 342/7106 (4.81%) |
| **Extremities** | **18/321 (5.60%)** | **799/7106 (11.24%)** |
| *Extremities sublocation\** |  |  |
| Hand | 4/321 (1.18%) | 60/7106 (0.84%) |
| **Trunk** | **11/321 (3.43%)** | **402/7106 (5.66%)** |
| *Trunk sublocation* |  |  |
| Chest | 4/321 (1.25%) | 211/7106 (2.97%) |
| Back | 9/321 (2.80%) | 445/7106 (6.26%) |
| **Shoulder** | **3/321 (0.93%)** | **243/7106 (3.42%)** |

\*Not all lesion sublocations are shown

Table 3 summarizes the different pathologic subtypes of high risk and non high risk BCC cohorts. Infiltrative (57.2%) and sclerosing (22.4%) were the most common subtypes for the high risk BCC group. The most common pathology subtypes for the non high risk BCC group were nodular (62.0%) and not otherwise specified (26.5%) histology.

**Table 3.** Pathology Subtypes^a^ of HR BCC and non-HR BCC.

| HR BCC |  |  |  |  | NON-HR BCC |  |  |  |
| --- | --- | --- | --- | --- | --- | --- | --- | --- |
| MN | SCL | INF | MOR | Total | NOS | SUP | NOD | Total |
| 59/339<br>(17.4%) | 76/339<br>(22.4%) | 194/339<br>(57.2%) | 10/339<br>(3.00%) | 339 | 2025/7655<br>(26.5%) | 887/7655<br>(11.5%) | 4743/7655<br>(62.0%) | 7655 |

<sup>a</sup>MN = Micronodular; SCL = Sclerosing; INF = Infiltrating; MOR = Morpheaform; NOS = Not otherwise specified; SUP = Superficial; NOD = Nodular.

Table 4 summarizes high risk and non-high risk BCC lesion diameter and depth. The mean diameter and depths of HR vs non-HR BCCs were similar with mean diameter slightly higher for the HR group(1.40cm) vs the non HR BCC subset (1.27cm). Mean depths recorded by high resolution dermal ultrasound (HRDUS) for both groups were equal(1.4mm each), however the median depth for non-HR cohort was lower (1.0 mm) vs the non HR cohort (1.35mm).

**Table 4.** BCC initial lesion sizes.

| BCC Subcategories | HR BCC | NON-HR BCC |
| --- | --- | --- |
| **Lesion diameter (cm) at start** | **N = 399** | **N = 7655** |
| Mean ± SD | 1.4 ± 1.00 | 1.27 ± 0.84 |
| Range | 0.3–8.0 | 0.0–10.5 |
| Median | 1.0 | 1.0 |
| **BCC lesion depth (mm) at start** | **n = 339** | **n = 7655** |
| Mean ± SD | 1.4 ± 1.00 | 1.4 ± 0.53 |
| Range | 0.3–8.0 | 0.0–4.82 |
| Median | 1.0 | 1.35 |

Table 5 illustrates the tumor stage (based on AJCC 8^th^ edition applied to all body locations rather than just the H&N region to be consistent) separated by high risk and non high risk BCC lesions. The HR BCC cohort had slightly higher proportion of stage II and III lesions whereas the non-HR cohort had a slightly higher proportion of stage I lesions (74% vs 80%).

**Table 5.** Stage of NMSC lesions.

|  | HR BCC | NON-HR BCC |
| --- | --- | --- |
| Stage | n (%) | n (%) |
| I (0 to <2 cm) | 252 (74.34) | 6157 (80.43) |
| II (2 to <4 cm) | 74 (21.83) | 1295 (16.92) |
| III (≥ 4 cm) | 11 (3.24) | 96 (1.25) |
| Unspecified | 2 (0.59) | 107 (1.40) |
| **Total** | **339(100)** | **7655 (100)** |

Table 6 summarizes the total fractions of treatments, total treatment dose, duration of treatment, and duration of follow up separated by the HR vs non-HR cohort. The total treatment fractions were the same amongst the 2 groups with a slightly higher total doses given to the HR BCC group (5502.4 cGy) vs the non-HR BCC group (5386.5 cGy). High risk lesions were treated for an average of 7.2 weeks and had an average follow up interval of 80.6 weeks. Non high risk lesions were treated for an average of 6.7 weeks with an average follow up interval of 101.17 weeks.

**Table 6.** Total number of treatment fractions, dose, duration, and follow-up interval.

| Characteristic | Statistic | HR BCC | NON-HR BCC |
| --- | --- | --- | --- |
| Total number of fractions | *N* | 339 | 7655 |
| | Mean ± SD | 20.3 ± 0.94 | 20.0 ± 1.24 |
|  | Range | 13.0 to 25.0 | 12.0 to 32.0 |
|  | Median | 20.0 | 20.0 |
| Total treatment dose (cGy) | *N* | 339 | 7655 |
| | Mean ± SD | 5502.4 ± 228.60 | 5386.5 ± 234.08 |
|  | Range | 4453.4 to 6595.88 | 3716.0 to 7363.7 |
|  | Median | 5491.2 | 5420.3 |
| Duration of treatment (weeks) | *N* | 339 | 7655 |
| | Mean ± SD | 7.2 ± 2.13 | 6.7 ± 4.17 |
|  | Range | 0.14–24.29 | 0.14 to 61.71 |
|  | Median | 6.86 | 6.57 |
| Duration of follow-up (weeks) | *N* | 339 | 7655 |
| | Mean ± SD | 80.6 ± 70.92 | 101.17 ± 89.17 |
|  | Range | 0.14–362.86 | 0.0 to 391.0 |
|  | Median | 58.9 | 77.14 |

Table 7 summarizes the energies that the lesions were treated with (50kV, 70kV, or 100 kV exclusively, or mixed) Lesions that were treated with more than one energy during the treatment course were termed as “mixed”. Approximately 3.5% of high risk lesions and 11.2% of non-high risk lesions were treated with a mix of two or more energies.

**Table 7.** NMSC lesion treatment by energy.

|  | HR BCC | Non-HR BCC |
| --- | --- | --- |
| Energy (kV) | N = 339 | N = 7642 |
| 50 | 21 (6.2%) | 1410 (18.5%) |
| 70 | 167 (49.3%) | 4059 (53.1%) |
| 100 | 139 (41.0%) | 1322 (17.3%) |
| Mixed | 12 (3.5%) | 851 (11.1%) |
| Total* | 339 (100%) | 7642 (100%) |

\*Some lesions did not have any lesion energy recorded

Table 8 highlights the safety profile of IGSRT treatment for both groups. Overall, in both groups combined approximately 4% (3.97%) of lesions had RTOG grade 3 or 4 toxicities showing the IGSRT treatment appeared safe and well tolerated and similar in both groups.

**Table 8.**
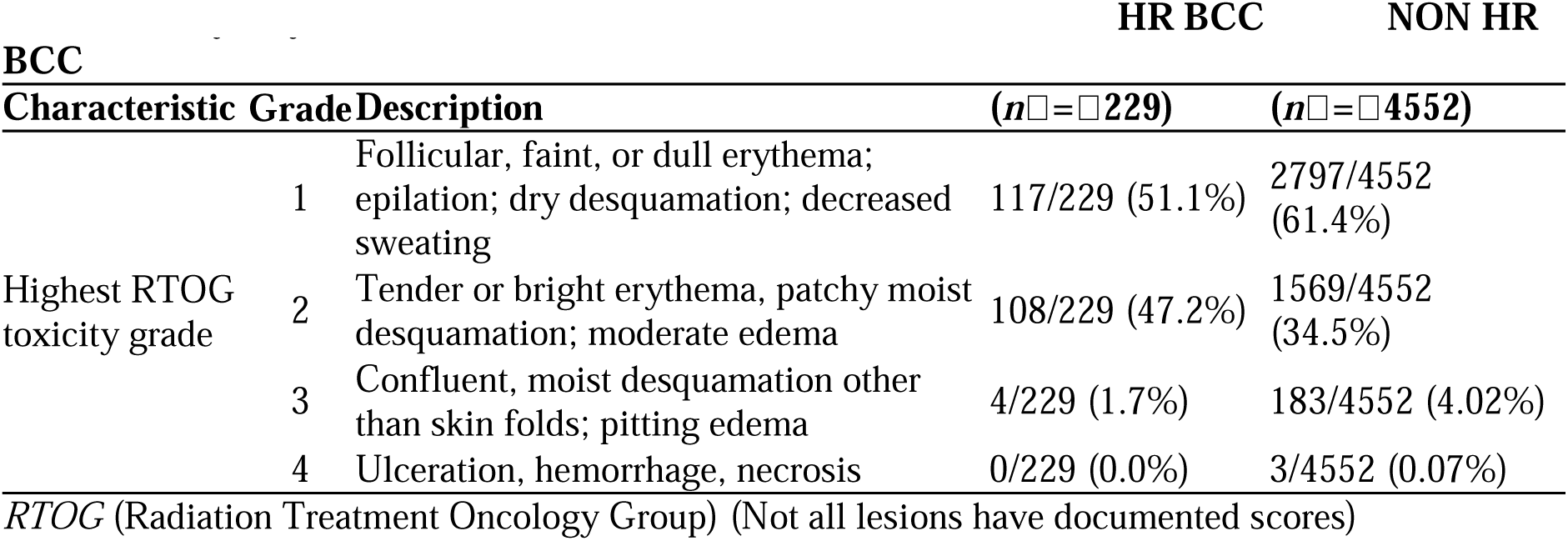
Safety—by lesion based on RTOG criteria.

### 3.1 Lesion Local Control

With a mean follow up of over 1.5 years in the HR cohort and almost 2 years in the non-HR group (going well past 5 years in a substantial number of lesions), there were no failures/recurrences in the HR BCC cohort w 100% local control. In the non-HR BCC cohort there were 31 failures.

#### Actuarial Local Control at 2 and 5 years

An actuarial 2 yr LC of 99.7% and 5 yr actuarial LC of 99.2% was achieved (Figure 1) in the non-HR group. The actuarial LC rates for the HR group were 100% at both 2 and 5 years (Figure 2). There was no statistical difference between the 2 cohorts’ actuarial LC rates on log-rank testing (p= 0.278) (Figure 3).

**Figure 1:**
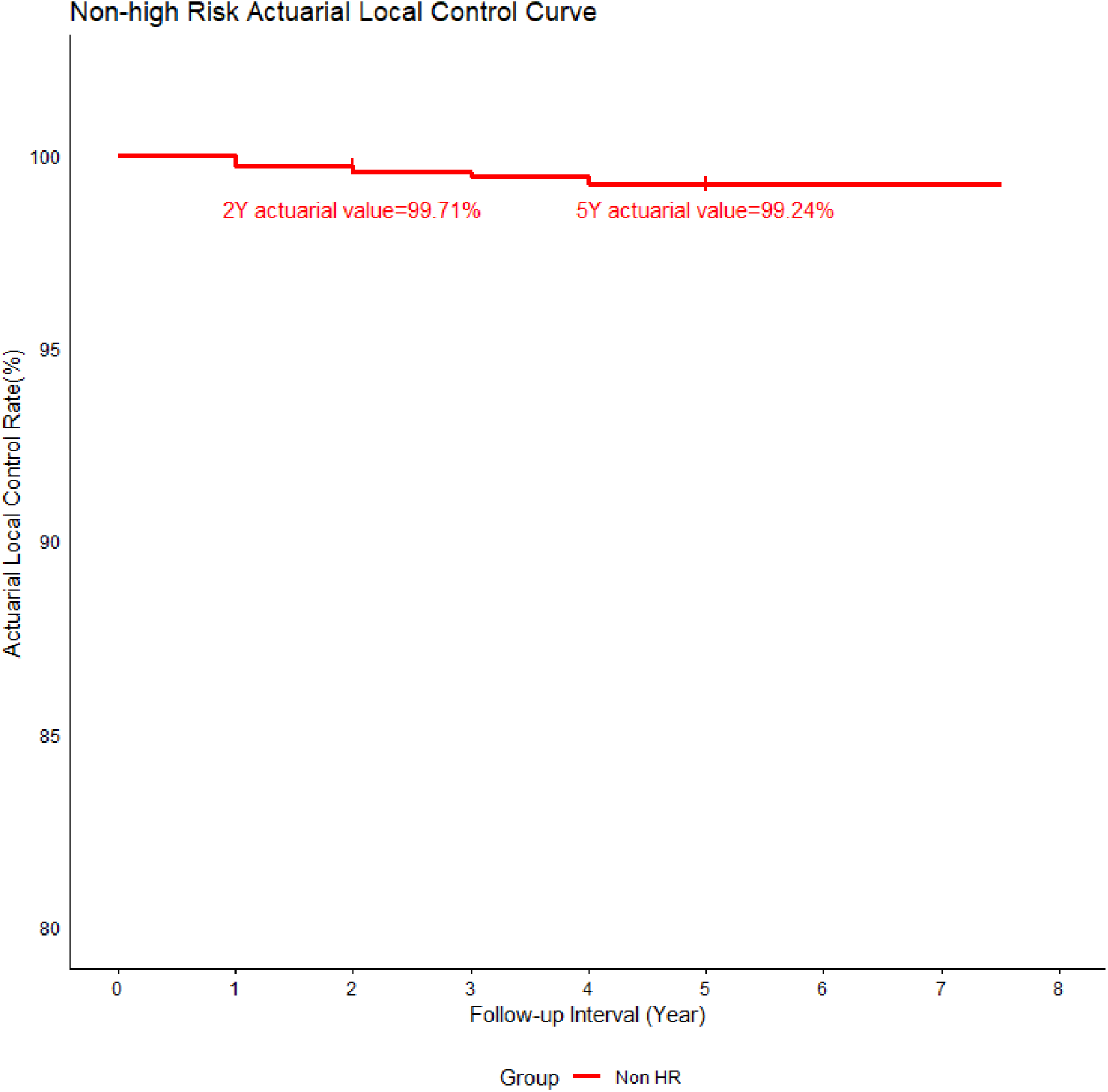
Actuarial Local Control at 2 and 5 Years for non-High Risk BCC. The graph demonstrates high rates of local control with actuarial 2 year local control of 99.71% and 5 year actuarial local control of 99.24%.

**Figure 2:**
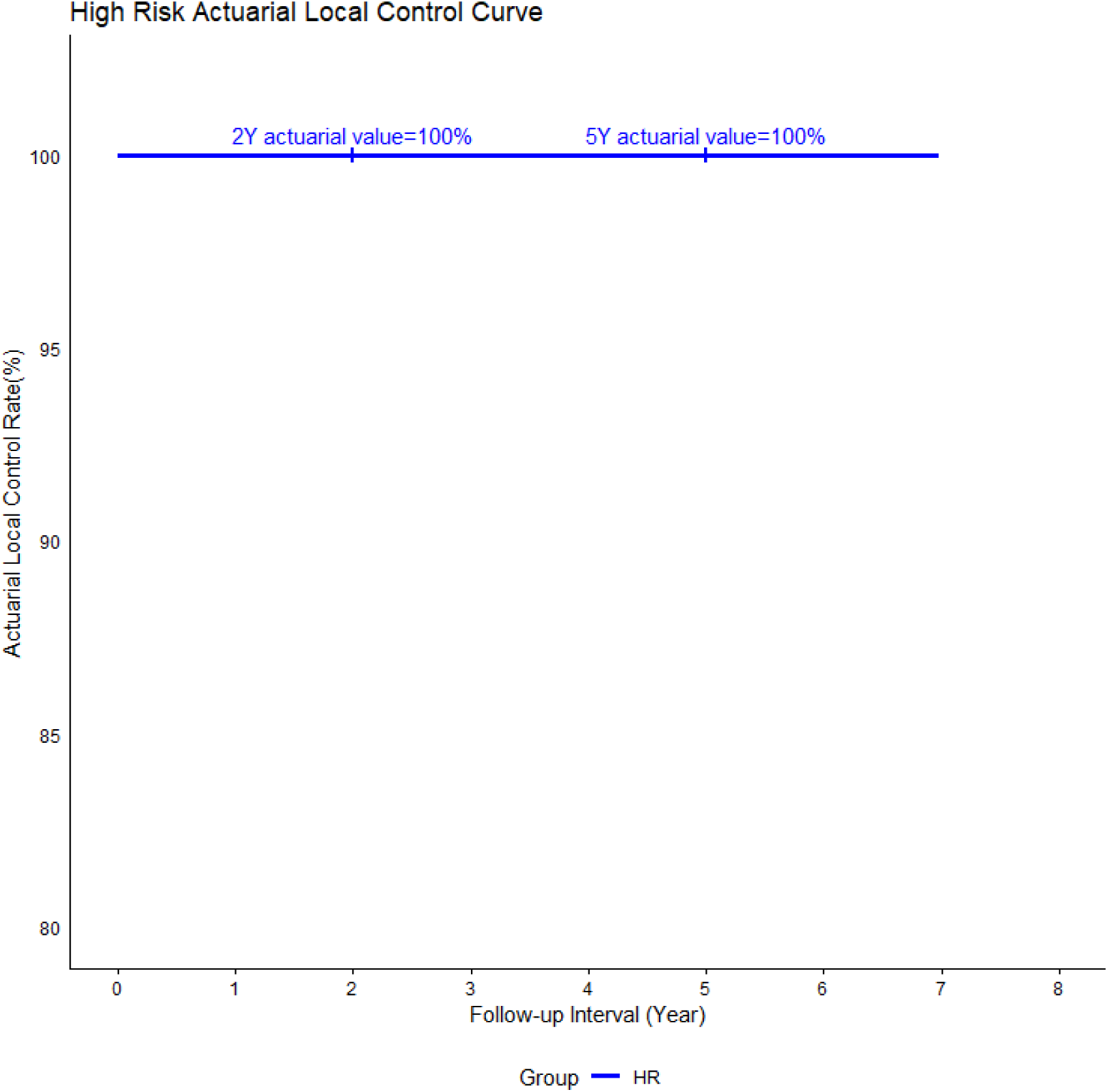
Actuarial Local Control at 2 and 5 Years for High Risk BCC. The graph demonstrates high rates of local control with actuarial 2 and 5 year local control of 100% and 100%.

**Figure 3:**
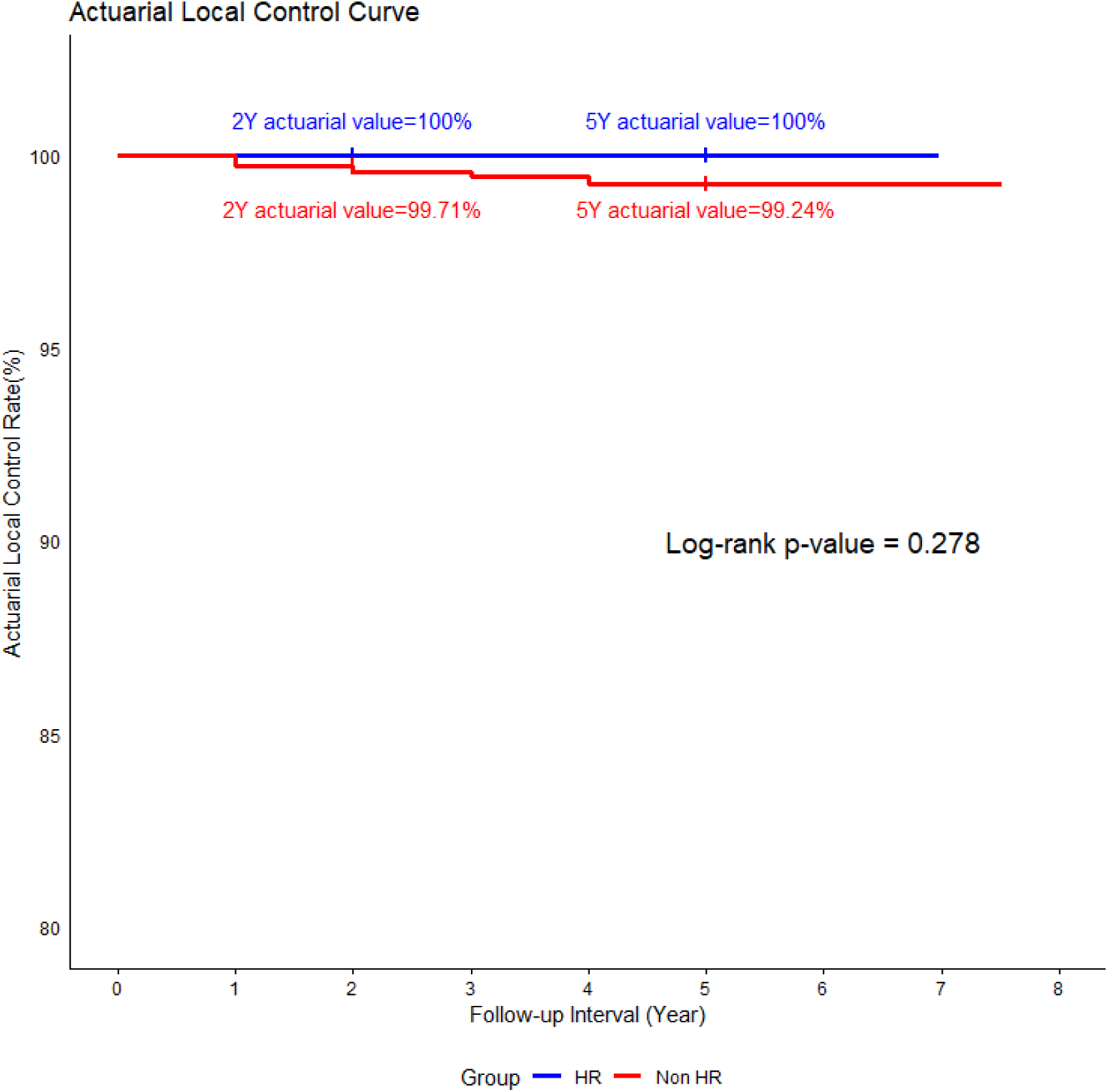
Actuarial Local Control Curve comparing High Risk and non-High Risk BCC groups. The blue line represents High Risk BCCs, and the red line represents non High Risk BCCs. The log rank test revealed a p value of 0.278 indicating no statistical difference in local control rates between the two groups.

#### Kaplan-Meier Local Control at 2 and 5 Years

The non-high risk 2 yr LC rate was 99.6%, and the 5 yr LC rate was 99.2% (Figure 4). The Kaplan-Meier analysis shows 2 and 5 yr LC of 100% (Figure 5) for the high risk subtypes. Statistical analysis using the log-rank test revealed no significant difference (p = 0.278) in local control rates between the two cohorts (Figure 6).

**Figure 4:**
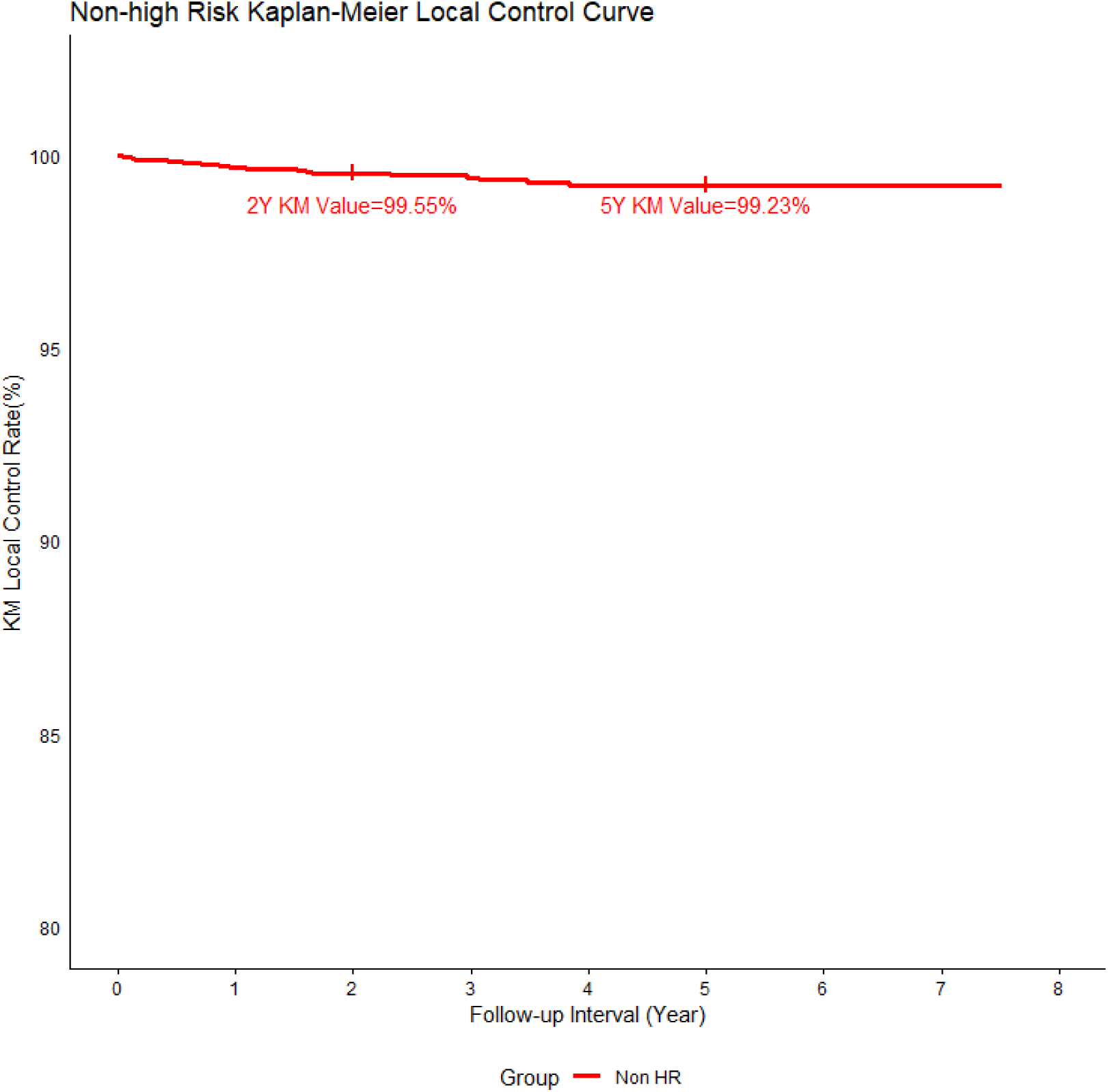
Kaplan-Meier Local Control at 2 and 5 Years for non-High Risk BCC. Kaplan-Meier local control curves for the non-High Risk BCC cohort at 2 and 5 years treated with IGSRT is 99.55% and 99.23%, respectively.

**Figure 5:**
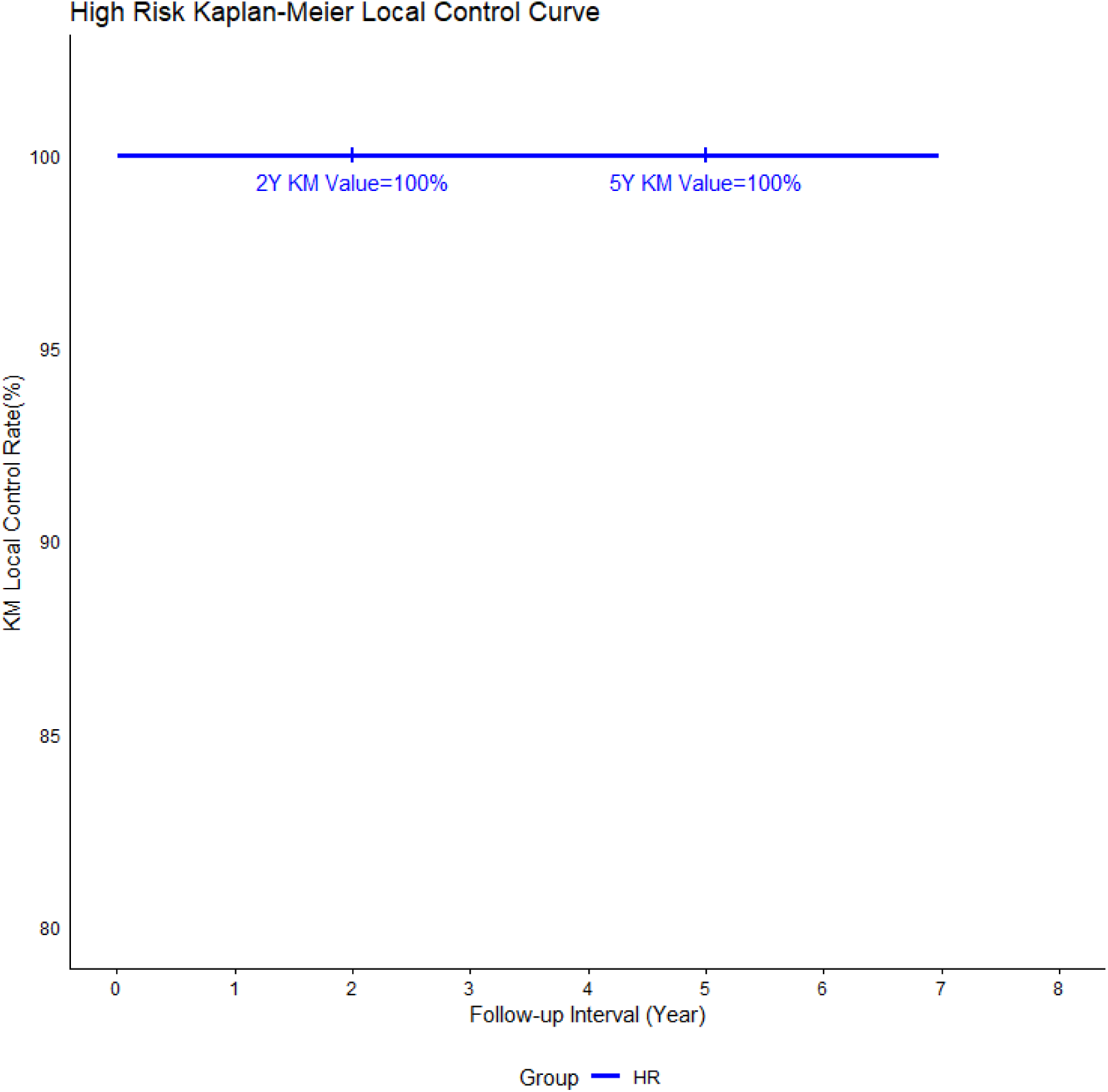
Kaplan-Meier Local Control Curve for High Risk BCC. The 2 year and 5 year KM local control rate using IGSRT are both 100%.

**Figure 6:**
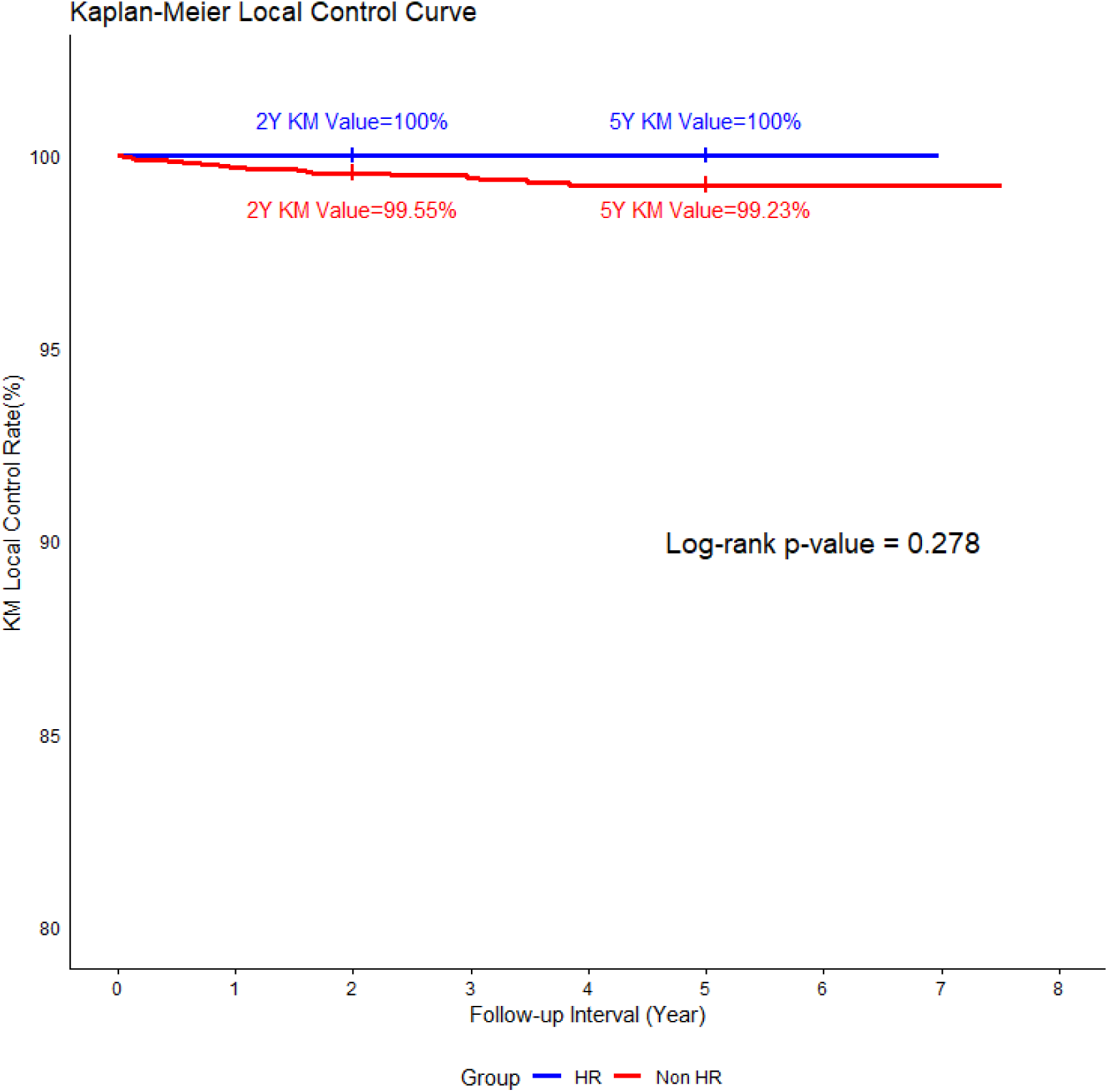
Kaplan-Meier (KM) Local Control (LC) at 2 and 5 years: KM Local Control Curves comparing High Risk and non-High Risk BCC groups. The blue line represents High Risk BCCs, and the red line represents non High Risk BCCs. The log rank test revealed a p value of 0.278 indicating no statistical difference in local control rates between the two groups.

## 4 Discussion

This paper presents a comprehensive analysis of superficial radiotherapy (ultrasound guided) treatment on the largest reported series of BCC patients in the United States, with one cohort pathologically diagnosed with high risk BCC subtypes consisting of 339 lesions of infiltrative, morpheaform, micronodular and sclerosing subtypes and the other non-high risk BCC cohort (nodular, superficial and NOS subtypes) comprising of 7655 lesions. Both cohorts underwent a similar treatment regimen with doses and energies using HRDUS guided superficial radiation therapy (commercially and commonly referred to as IGSRT) using a previously published regimen (Ladd-Yu protocol) [12].

The HR BCC cohort showed 100% KMLC which was minimally ( and non statistically) higher than the non-HR BCC subgroup (99.6% and 99.2% KM LC) at 2 and 5 years, respectively.

Both HR and non HR groups had low toxicity of treatment with the regimen used showing RTOG toxicity generally limited to grades of 1 and 2, and rare instances of RTOG 3 and 4 toxicity.

These results shows that high risk histology subtypes of BCC are indeed radiosensitive using SRT combined w HRDUS guidance. In fact, these aggressive BCC subtypes may actually be paradoxically more radiosensitive when compared to standard/non-aggressive BCC subtypes. Although the number of HR cases are lower than the non-HR BCC group, the authors feel that there are substantially sufficient number to justify the validity the results and conclusions. This observation may actually be consistent with certain other higher grade cancers subtype radiation response showing a similar paradoxically higher radiosensitivity and initial complete response rate than lower grade cells such as in Bladder CA and lymphomas, etc [16]. Albeit in these other decidedly more life threatening cancers with their tendency for distant or nodal metastases, the initial response may not translate to better outcome. We feel that for the typically local only behavior of BCCs, that the radiosensitivity and complete response rate does indeed translate to ultimate long term local control or “cure” (a term many dermatologists like to use for BCCs and other early stage NMSC) of the target lesion.

The LC differences between the 2 groups are small(<1%) by log rank analyses with both cohorts showing outstanding LC out past 5 years and no statistically significant difference in lesion local control rates by log rank analysis. This improved LC in all subtypes of BCCs establishes that the relatively new modality of IGSRT is appropriate for all BCCs regardless of subtype.

### Debunking Myths of radioresistence of HR BCC

These results contribute to debunking the myth that radiation therapy is ineffective or less effective for high risk BCCs. Our analysis shows an equivalent or actually lower failure rate (of 0%) compared to the non-high risk cohort (at 0.5-0.8% failure).

The most obvious explanation for this improvement is the new ability of IGSRT over tradiational non-image guided radiotherapy modalities. In 2 recent studies using logistic-regression analysis and meta-analysis, LC in the treatment of NMSC using IGSRT in comparison to non-image guided external radiation modalities (superficial radiation therapy (SRT) or external beam radiation therapy (XRT including electron beam or photon beam radiotherapy) have been shown to be superior and achieving statistically improved LC [14,15] and were attributed to the image guidance factor. Whereas previously, the use of RT may not have been recommended for high risk BCCs due to potentially higher recurrence rates, the advent of HRDUS in combination with SRT allow more precision to be directed to the full depth of the tumor and ushers in an era of modern more exacting targeting of radiation beams that appears to be associated with higher success rates rivaling that of surgical interventions such as MMS and present a valuable treatment option especially for those who are not ideal candidates for surgical management. This paradigm shifting development uses the HRUS which has high correlation with depth of tumor involvement [17] and is useful not only in pre-treatment planning but also in evaluation during the midst of treatment as well as for following the lesion post treatment at the post therapy area in both scenarios of surgical and non-surgical (including definitive radiotherapeutic) treatment approaches. The imaging can give information to help in treatment modification/adaptation and direct potential salvage therapy selection post therapy. HRDUS may also be helpful to help differentiate the aggressive subtypes of BCC versus non-aggressive subtypes based on the echogenicity of the ultrasound findings [17]. Further discussion of the use of HRUS is beyond the scope of this paper and may be a good subject for future publications. The utility of HRDUS in surgical NMSC scenarios may benefit from further research endeavors.

Based on the results of this study, IGSRT may indeed be a good option in these HR subtypes and in select situations may be preferred over surgical excision/MMS in situations for patients with surgical fatigue, lesions in cosmetically/functionally sensitive locations, poor performance status subjects on anticoagulation, or those who cannot tolerate a prolonged surgical procedures requiring patient to remain still for extended periods of time, or those who decline surgery.

By examining the outcomes of these cases, this paper challenges the prevailing notions about the management of high risk and non-risk BCCs and provides evidence that may change treatment perspectives and future guidelines. A paradigm shift may be advocated for how high risk BCC is approached, moving away from previous notions of surgical only treatments to a more customized evidence-based approach that is also non-surgical.

As the prevalence of skin cancer continues to expand, such advancements are critical in the evolving landscape of cancer care ensuring that patients, in consultation with their dermatology and/or radiation/ oncology providers, make the best decision with regard to selection of the treatment modalities best suited for the patients’ specific situation to maximize optimal outcomes as well as assuring safety minimizing short term and long term toxicity and subsequently achieving the best quality of life, both functionally and cosmetically.

### 4.1 Limitations

A number of practices may have excluded these high risk BCCs categorically and steered these patients to Mohs after discussion. Others did not exclude any histological subtypes. The category of not otherwise specified BCCs may have certain BCCs that may have elements of high risk features that were simply not reported and perhaps might have accounted for some of its failures.

Competing risks of death was not accounted for however, we feel that BCC typically does not affect mortality, thus we felt exclusions based on this was not necessary and thus was not done in this paper.

## 5 Conclusion

IGSRT achieves high success rates in all subtypes of BCC in retrospective review on almost 8,000 cases in the USA.

HR BCC subtypes may actually be paradoxically more radiosensitive than or at least equivalent to other more common “non-aggressive” BCC subtypes.

There is no contraindications to using RT in the form of SRT combined w HRDUS (commonly called IGSRT) for high risk subtypes of BCCs. In fact, in certain patients with any subtype of BCC, IGSRT may be preferential over excision/MMS if the patient is a poor surgical candidate, has lesions in cosmetically/functionally sensitive locations or if the patient prefers a non-surgical approach.

## Data Availability

III. Availability of data and material:
The data underlying this article will be shared on reasonable request to the corresponding author.

## Declarations

### III. Availability of data and material

The data underlying this article will be shared on reasonable request to the corresponding author.

### V. Consent to participate

Not applicable

### VI. Consent for publication

Not applicable

### VII. Code availability

Not applicable

### VIII. Author contributions

All authors have contributed to the manuscript and approved the final submitted version of the manuscript. Lio Yu-Conceptualization, methodology, data collection and curation, supervision, data and statistics verification, writing-original draft, review and editing. Michael Kaczmarski-data collection, data and statistics verification, tables and figures, writing-original draft, review and editing. Clay J. Cockerell-concept refinement, supervision, review and editing.

## Acknowledgements

The authors would like to acknowledge the following people who were instrumental in collecting and extracting data from various different locations, without whom this study would not be possible:□Amanda Boatner, Berta Benzschawel, Beth Ann Marogi, Brianna Moore, Carolyn Mahoot, Charles Bingner, Cherie Hoppe, Diane Krause, Lio Yu, Eric Hill, Erica Kovaluskie, Isabella Moore, Jacob Britt, Jacqueline Contino, Jennilee Kovaluskie, Jenny Folden, JoAnne Porta, Karen McKenna, Katie Curtiss, Kim McLaugherty, Kimberly Wilke-Zuma, Krupal Patel, Lacy Smith, Leslie Walker, Lisa Gonzalez, Lisha Kelly, Macie Najera, Mairead Moloney, Mandi Brown, Michael Kaczmarski, Michelle Smith, Nick Natalizio, Olivia Desalvo, Paula Wright, Peter Kaczmarski, Prithvi Narasimhan, Rachel Lindley, Rachelle Moore, Samantha Sheehan, Sarah Moss, Soham More, Stacey Robinson, Tonya Johnson, and Yancy Boatner. Special thanks to Soham Moore and Prithi Narasimhan from Sympto Health for data scraping of data from EMR’s. We are indebted to Songzhu for statistical calculations and LC curve generation.

